# The *Milieu Intérieur* follow-up study – an integrative approach for studying immunological variation in an aging population

**DOI:** 10.1101/2024.09.17.24313506

**Authors:** Anthony Jaquaniello, Florian Dubois, Farah Rahal, Tom Dott, Slobodan Culina, Chloe Albert-Vega, Vincent Rouilly, Etienne Villain, Violaine Saint-André, Divya Unni, Sandrine Fernandes Pellerin, Emma Bloch, Michael White, Milena Hasan, Lluis Quintana-Murci, Etienne Patin, Darragh Duffy, Milieu Intérieur Consortium

**Author notes:** Equal contributions (Florian Dubois, Farah Rahal, Tom Dott, Slobodan Culina, Chloe Albert-Vega). Equal contributions (Lluis Quintana-Murci, Etienne Patin, Darragh Duffy).

## Abstract

Human immune responses are highly variable from one individual to another, with these differences determined by both genetic and environmental factors. We previously established the Milieu Intérieur cohort to define the boundaries of healthy immune responses and identify and quantify its different determinants. From this cohort of 1,000 well-defined healthy donors, we have identified multiple genetic and environmental factors that are associated with variable immune phenotypes. Crucially, these associations were observed in a healthy context, as defined by strict inclusion and exclusion criteria. To assess how such immune responses may vary over time, we initiated a 10-year follow-up clinical study of the cohort. From the original 1,000 donors, we identified and recruited 415 donors from whom we assessed recent medical history and lifestyle factors, in addition to collecting samples for measurement of diverse immune phenotypes. A comparison of 107 laboratory and serological measures over the 10-year period showed significant correlations, highlighting the quality of the data collected. By accounting for potential batch effects, our analyses revealed the profound effects of aging on health-related biomarkers and antibody levels against common pathogens. We determined that a significant proportion of the donors no longer meet the initial definition of good health, due to either physiological changes associated with aging, or disease incidence including cancer, autoimmune and cardiovascular diseases. We found that disease development over the past 10 years is associated with a biological aging score, assessed at the time of the initial recruitment. In summary, the Milieu Intérieur follow-up study will provide new opportunities for studying biological mechanisms of immunosenescence and identifying factors associated with accelerated immune aging and poor health outcomes.

## Introduction

Individuals exhibit substantial variation in terms of disease risk, response to treatments, and clinical outcomes. These differences may arise from naturally-occurring variability in immune responses, which can be due to factors such as age, sex, gender, genetic background, socio-economic status, and environmental exposures^1,2^. Age, in particular, appears to have one of the strongest influences, affecting basic physiological processes through biological aging and changing environmental exposures over time^3–5^. While an increasing number of studies have examined age-related differences in immune responses^6–10^, few have investigated the molecular and cellular mechanisms underlying human immune aging in natural settings^11,12^. The Stanford longitudinal aging study reported important, significant inter-individual differences in aging immune responses^13^, but the factors contributing to this phenomenon remain unclear.

To identify the determinants of immune variability, the Milieu Intérieur (MI) population cohort study^14^ was established in 2012-2013, consisting of 1,000 well-defined healthy donors equally stratified by age and sex (100 male and 100 female donors in each decade of life between 20-69 years old). From these donors, an extensive biocollection of skin, nasal, fecal, and baseline and stimulated blood samples was established, along with comprehensive health and lifestyle information. An ever-growing extensive data warehouse was established, encompassing genomic^15^, epigenomic^16^, transcriptomic^17^, proteomic^18,19^ cellular^15^, serological^20^, and microbiome^21^ datasets for the entire cohort. Using this rich and unique dataset, MI has so far identified and quantified the impact of genetic variants, age, sex, smoking, socioeconomic status, persistent infections, blood group antigens, BMI, and diet on variability in immune cell composition^15^ thymic function^22^, circulating proteins^18^, and functional immune responses at both baseline levels^20^ and after stimulation with microbes, agonists, and cytokines^17,23,24,25,26^.

To address our original questions on understanding immune variability, the MI study was designed as a cross-sectional study with a narrow time frame (with half of the donors resampled within 2-6 weeks), a focus on healthy individuals, and a defined age range (20-69 years of age)^14^. However, such a design precludes the assessment of immune variation with time and beyond 70 years of age, the study of the impact of disease incidence on immunity, and the role of age-related immune perturbations in disease development. We recently addressed these limitations through a longitudinal assessment of available donors 10 years after the original study. As the study was not initially designed for longitudinal assessment, anticipating donor participation was challenging. However, following a broad information campaign, we successfully recruited 415 of the original 1,000 donors. As we wanted to study disease incidence and outcomes, exclusion criteria were minimal. The same information and biological samples were collected during the new sampling campaign for cross-comparison over the 10-year period. Additionally, given the onset of the COVID-19 pandemic in 2020, we took advantage of this follow-up study to assess the biological underpinnings of immune responses against SARS-CoV-2, by expanding our clinical investigation to coronavirus infections and assessing antibody titers against common coronaviruses at both time-points. Here, we present a description of the 10-year follow-up study, assess how the biochemical, serological, and clinical lab measures have changed through time, and evaluate how these biomarkers predict the health incidences observed in this well-characterized aging cohort.

## Material and Methods

### Clinical protocol and implementation

The 10-year follow-up Milieu Intérieur study, referred to as MI visit 3 (“V3”), was approved by the Comité de Protection des Personnes — Nord Ouest III (Committee for the protection of persons) on 27th January 2022, and by the French Agence nationale de sécurité du médicament (ANSM) on 30th November 2011. The study was sponsored by the Institut Pasteur (ID-RCB Number: 2021-A02621-40) and conducted as a single center study without any investigational product. The protocol is registered at ClinicalTrials.gov (study# NCT05381857). As this study was designed to be a 10-year follow-up of the original Milieu Intérieur “V1” cohort (NCT01699893 and NCT03905993), the major inclusion criterion was previous inclusion in the V1 study. In addition, donors were required to give written informed consent, and be affiliated to the French social security regimen. Non-inclusion criteria for these pre-screened individuals were restricted to ongoing pregnancy, inability to provide informed consent, or specific reasons requiring legal protective measures. No donors were excluded for any of these non-inclusion criteria. In total, 415 subjects from the previous Milieu Intérieur cohort were included. All subjects received compensation for their participation.

### Donor recruitment

As for the original Milieu Intérieur study, donors were recruited at Biotrial, Rennes, France (Contract Research Organization, SIREN: 351 643 523). Donors were first informed of the study through an information leaflet, which invited them to participate in an online webinar. During the webinar, results from the first study were presented and participants had opportunities to ask questions to scientists from the consortium (https://www.milieuinterieur.fr/en/contact-us/webinar/). In the second stage, donors were contacted directly through invitation letters, emails, and telephone calls.

### Biological sampling

Donors were interviewed and sampled during a single visit at Biotrial (Rennes, France) between the period of 14^th^ March 2022 and 12^th^ October 2022. After completing a RedCap-based healthcare and lifestyle questionnaire (Supplementary Text) with a medical practitioner, all donors underwent a complete medical check, as originally described^14^, and provided 100mL of blood and two nasal swab samples (**Table 1**). Fecal samples were collected at home by the donors in the days prior to the medical visit in Omnigene Gut OMR-200 tubes, which were frozen at −80°C upon reception at the clinic. Whole blood was used for routine clinical and biochemical lab testing (21 mL) (**Table 1**), as well as immediate incubation in TruCulture whole blood assays for 21 different immune ligands (25mL each), and 54mL was transported on the same day under temperature monitoring to the Institut Pasteur, Paris for flow cytometry analysis and peripheral blood mononuclear cells (PBMCs), plasma, and DNA isolation.

**Table 1.** Sample collections obtained from Milieu Intérieur donors. CLT, clinical laboratory test; RBC, red blood cell; HCT, hematocrit; HGB, hemoglobin; MCV, mean corpuscular volume; MCH, mean corpuscular hemoglobin; WBC, white blood cell, NEUTRO, neutrophil; LYMPHO, lymphocyte; EOS, eosinophil; BASO, basophil; PLT, platelet; Na, sodium; K, potassium; Ca, calcium; P, phosphorus; Cl, chloride; HCO3, bicarbonate; HSA, human serum albumin; ALP, alkaline phosphate; AST, aspartate aminotransferase; ALT, alanine aminotransferase; GGT, gamma-glutamyl transpeptidase; BILI, bilirubin; TPROT, total protein; CRP, C-reactive protein; BUN, blood urea nitrogen; CREAT, creatinine; UA, urinalysis; GLUC, glucose; TCHOL, total cholesterol; LDL, low density lipoprotein; HDL, high density lipoprotein; TRIGLY, triglycerides; βHCG, beta-human chorionic gonadotropin. TBD, to be done.

| Data type | Biological measures | Analysis site |
| --- | --- | --- |
| **Whole blood collection (*n* = 411)** |  |  |
| Complete blood count | RBC, HCT, HGB, MCV, MCH, WBC, NEUTRO, MONO, LYMPHO, EOS, BASO, PLT | Biotrial |
| Blood electrolytes | Na, K, Ca, P, Cl, HCO<sub>3</sub> | Biotrial |
| Liver function tests | HSA, ALP, AST, ALT, GGT, BILI, TPROT | Biotrial |
| Inflammation | CRP | Biotrial |
| Renal function tests | BUN, CREAT, UA | Biotrial |
| Lipids/metabolism | GLUC, TCHOL, LDL, HDL, TRIGLY | Biotrial |
| Antibody subtype levels:<br>Immunoglobulin electrophoresis | Total IgM, IgG, IgA, IgE serum concentrations | Biotrial |
| Microbial specific antibody levels:<br>ELISA assays | Hepatitis B (HBs Ag), Hepatitis C (anti-HCV IgG), HIV (anti-HIV IgG, anti-HIV IgM), CMV (anti-CMV IgG), HTLV-1 (anti-HTLV-1 IgG), Influenza | Biotrial |
| Microbial specific antibody levels:<br>Luminex assays | Immunoglobulin concentrations against 72 microbial antigens | Institut Pasteur |
| Blood immune cell composition and immunophenotyping:<br>cytometric analysis (Na Heparin tube) | TBD | Institut Pasteur |
| Functional immune stimulation (Na Heparin syringe): TruCulture tubes for 21 different immune stimuli | TBD | Stored at -80°<br>for later analysis |
| Genetic variation: DNA sequencing (EDTA tube) | TBD | Stored at -80°<br>for later analysis |
| Peripheral Blood Mononuclear Cells (PBMCs) (Heparin tube) | TBD | Stored at -80°<br>for later analysis |
| Plasma (EDTA tube) | TBD | Stored at -80°<br>for later analysis |
| **Urine collection (*n* = 414)** |  |  |
| CLT Biochemistry | proteinuria, glycosuria (dipstick) | Biotrial |
| CLT Pregnancy test | βHCG concentration (women only) | Biotrial |

| **Fecal sample collection (n=414)** |  |  |
| --- | --- | --- |
| OMNIgene GUT tube (OMR-200) | TBD | Stored at -80°<br>for later analysis |
| OMNImet GUT (ME-200) | TBD | Stored at -80°<br>for later analysis |
| **Nasal swabs (n=414)** |  |  |
| Copan FLOQ Swabs (left and right nostril) | TBD | Stored at -80°<br>for later analysis |

### Luminex-based serotyping

Serum samples were tested for antibodies to a broad panel of common respiratory pathogens and routine vaccine-preventable diseases using bead-based multiplex assays. A 61-plex assay was developed to include antigens for adenovirus, cytomegalovirus (CMV), Epstein-Barr virus (EBV), echovirus, enterovirus CoxB3, Hepatitis A virus (HAV), Hepatitis B virus (HCV), Hepatitis C virus (HBV), measles, Varicella-Zoster virus (VZV), mumps, rubella, norovirus, respiratory syncytial virus (RSV), rhinovirus, rotavirus, human papillomavirus, and influenza A virus (IAV). In parallel, a 24-plex assay was developed for SARS-CoV-2 antigens and human seasonal coronaviruses including 229E, NL63, OC43, and HKU1. Development of these assays, including the antigens used, are described elsewhere^27^. The proteins used were either purchased from Native Antigen (Oxford, UK), ProSpec-Tany Techno Gene (Israel), or Ray Biotech (Georgia, US). Samples were run at a final dilution of 1:200. Plates were read using a Luminex IntelliFlex system, and the median fluorescence intensity (MFI) was used for analysis. Samples from V1 and V3 visits were randomized in the same batch. We discarded antibody levels for which seropositivity could not be defined unambiguously, showing evidence of cross-reactivity or poor reproducibility^27^. After quality control assessment, 72 serological measures were retained for subsequent analysis.

### Clinical data collection

Clinical data were collected and managed using REDCap electronic data capture tools^25^ (Research Electronic Data Capture v11.4.4) hosted at Institut Pasteur (Paris, France). REDCap is a secure, web-based software platform designed to support data capture for research studies, providing (i) an intuitive interface for validated data capture; (ii) audit trails for tracking data manipulation and export procedures; (iii) automated export procedures for seamless data downloads to common statistical packages; and (iv) procedures for data integration and interoperability with external sources^28^. Data were collected by Biotrial between 14^th^ March 2022 and 28^th^ October 2022. The electronic clinical report form (eCRF) included a total of 1,050 questions and was divided across ten thematic forms: general information, medical history, cancers, vaccinations, infectious diseases, COVID-19, other information, food habits, biological samples, and adverse events. The questionnaire, written in French, is provided as Supplementary Text.

### Clinical data cleaning

After clinical data collection, each form was exported from REDCap separately and imported in the R software (v. 4.3.0). Data cleaning comprised various steps: standardization of the variable names, formatting of the date variables to YYYY-MM-DD ISO-8601 format, English translation and uniformization of the ‘free text’ variables with regular expressions. Several quality controls were performed; including the detection of age and sex discrepancies, and the detection of outlier values in the clinical data, defined as values outside of expected boundaries provided by Biotrial. Each discrepancy/outlier value was reported to Biotrial and corrected directly in REDCap, whenever possible.

### Data processing

Two donors presented with poor venous capital and one showed evidence for hemolysis, which is known to impact the quality of laboratory measurements. These donors were excluded from biological analyses. One additional donor was excluded because of ongoing breast-feeding. In total, 411 out of 415 donors were kept for biological analysis, with 415 used for questionnaire-based analysis. For all V1 and V3 biological and serological quantitative measurements, only three data points were missing and were imputed with the K-Nearest-Neighbors (K = 5) imputer (scikit-learn package v1.4.1)^26^. Eleven outlier values, defined as physiologically unlikely data points out of the interval bounded by the first quartile – 4.5 × interquartile range (IQR) and the third quartile + 4.5 × IQR, were replaced by the median of the distribution. For specific analyses, variables were either scaled between 0 and 1 by min-max scaling or converted into *Z*-scores by standardization, to facilitate comparisons across variables. All the processing steps were performed in Python 3.10.4 with packages numpy^29^ 1.26.4, pandas 2.2.1, scikit-learn 1.4.1, in a Jupyter-lab 3.4.2 web-based notebook.

### Statistical analyses and data visualization

Statistical tests and multiple regression analyses were conducted in Python 3.10.4, with packages scipy 1.12.0, statsmodel 0.14.0. R software 4.3.0 was also used for Generalized Linear Models (GLM) and Generalized Linear Mixed Models (GLMM) with R packages lmerTest^30^ 3.1-3 and lme4^31^ 1.1-35.1. To assess non-linear effects of age, spline regressions and Likelihood-ratio tests were performed with R packages splines 4.3.0 and lmtest 0.9-40. Tests were corrected for multiple testing with the Bonferroni correction. Candidate risk factors for SARS-CoV-2 infection and COVID-19 severity included age, sex, smoking status, BMI, abdominal circumference, blood pressure, heart rate and C-reactive protein concentration^32^. The SARS-CoV-2 infection severity score was computed as previously proposed^33^, based on self-declared symptoms, treatments and hospitalization. PhenoAge was computed with the R package BioAge^34^ 0.1.0. As the red cell distribution width (RDW) variable was missing from the MI data, we trained a new PhenoAge model on the NHANES III data without the RDW predictor. The metabolic score, which varies from 0 to 1, was computed for each donor by incrementing the score by 1 when: abdominal circumference > 94 cm or > 80 cm in men or women, respectively; systolic blood pressure ≥ 130 mmHg or diastolic blood pressure ≥ 85 mmHg; triglyceride levels ≥ 1.7 mM; HDL levels < 1 mM or < 1.3 mM in men or women, respectively; glucose concentration ≥ 6.1 mM^35^. Data visualization was performed using Python package Matplotlib 3.8.0 and Seaborn 0.13.2, or R package ggplot2 3.4.3 and Patchwork 1.2.0. A conda environment recipe file in yaml format with the specified package versions is available upon request, to rebuild the environment and reproduce the results.

## Results

### The Milieu Intérieur V3 cohort

We initially established the Milieu Intérieur (MI) cohort in 2012-2013 to evaluate how sex, age, and environmental and genetic factors affect inter-individual variation in immune responses. A total of 1,000 healthy donors were recruited in western France, with an equal number of females and males (*n* = 500 of each sex) and participants from five decades of age^14^ (20-69 years, *n* = 200 per decade; Fig. 1A,B). The recruitment was designed to maximize the statistical power to detect effects of sex and age, as well as their interactions, on immunity. In 2021, we initiated a follow-up study of MI participants, referred to as “V3”, 10 years after the first sampling visit “V1” (Fig. 1A), to assess aging effects on immune responses beyond 70 years of age, to study inter-individual variability in immune aging, and to observe whether MI participants have developed diseases. We aimed to recruit the maximum number of donors from the original cohort and contacted all V1 donors by mail and phone, resulting in the recruitment of 415 participants.

**Figure 1.**
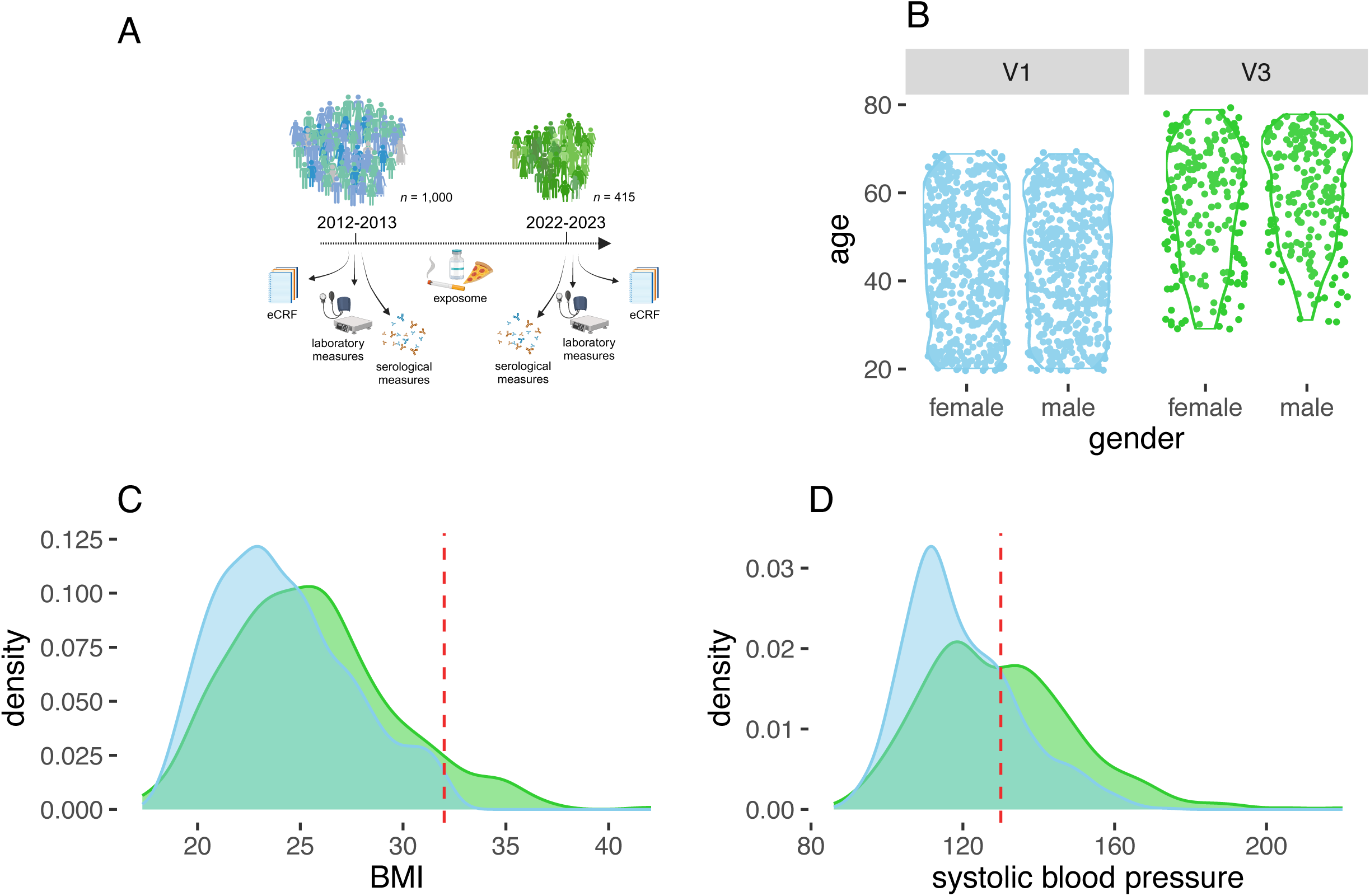
A 10-year follow-up study of the Milieu Intérieur aging cohort. (A) Sampling strategy for the Milieu Intérieur (MI) cohort. (B) Age distribution of MI donors at V1 and V3 visits, according to sex. (C) Body mass index (BMI, in kg/m²) density distribution at V1 and V3 visits. The red dashed line indicates BMI = 32 kg/m², the threshold value used as an inclusion criterion in V1. (D) Systolic blood pressure (mmHg) density distribution at V1 and V3 visits. The red dashed line indicates SYSBP = 130 mmHg, the value above which SYSBP is considered high^35^. (n=1000 (V1), n=415 (V3))

The follow-up V3 cohort was not biased according to sex and the sex ratio was not different from that of the V1 cohort (*n* = 211 and 204 females and male donors, respectively; chi-squared test *P* = 0.77). By contrast, there was a bias for age, with older donors more likely to participate in the new study, as shown by significant age differences between V1 donors who returned to participate in the V3 study compared to those who did not (chi-squared test *P* = 1.5 × 10^−16^; Fig. 1B). We next explored potential recruitment bias due to socioeconomic status (SES), using our recently described ELO rating system based on three variables: education, income, and home ownership^24^. We found no significant differences in ELO SES score between V1 donors who participated in the V3 study and those who did not (LRT *P* = 0.65), while controlling for age. Furthermore, we observed no significant associations between recruitment date (70 dates; *n* = 5.93 samples per date) and age (Kruskal-Wallis test *P* = 0.35) or sex (chi-squared test *P* = 0.73), nor between clinical investigator (18 investigators; *n* = 23 samples per investigator) and age (*P* = 0.24) or sex (*P* = 0.10), indicating that age or sex effects on biological measures are not expected to be confounded by potential batch effects.

For the 415 donors, we collected a wide range of demographic data through structured questionnaires (Supplementary Text), covering health-related habits (physical activity, nutritional habits, sleep quality, etc.) and medical and vaccination records. We also collected 45 biological measurements, including morphological data (e.g., height, weight), general health parameters (e.g., blood pressure, heart rate, temperature), and health biomarkers (e.g., lipid plasma levels, blood cell counts, proteinuria) (Table 1). As expected, we observed that 221 V3 donors no longer met all of the strict inclusion criteria used in V1, suggesting that they had become less healthy with age. For example, V3 participants have, on average, higher BMI and blood pressure than V1 participants (Fig. 1C,D). Together, these analyses indicate that the MI follow-up study is well designed to test how aging and age-related common health conditions affect the immune system.

### Biological measurements in an aging cohort

We evaluated the quality of the newly collected data by computing, for each donor, the correlation between biological measurements collected at V1 and V3 visits. To identify potential mismatched pairs of samples, we compared paired correlations to unpaired correlations computed for all donors. We found six donors with unexpectedly low correlations (Spearman’s *r* < 0.32 at FDR = 5%; Figs. 2 and S1), suggesting these V1 and V3 biological data were not collected from the same donor. Although genetic data will be needed to confirm these observations, conservatively we removed the six donors from subsequent analyses. We then computed correlations for each biological measurement between 405 V1 and V3 donors with complete data, to identify variables with high measurement error. We found two biological measurements, chlorine and potassium levels, that showed lower V1/V3 correlations than expected (Spearman’s *r* < 0.31 at FDR = 5%; Figs. 2B and S2), relative to other paired correlations. Additionally, we estimated the effect of data sampling batches on the biological measurements, by testing for differences in biological measurements among sampling days or clinical investigators. Out of the 45 variables tested, we found that chlorine (LRT *P* = 5.4 × 10^−13^; Fig. S3), sodium (LRT *P* = 1.3 × 10^−8^), and albumin (LRT *P* = 7.4 × 10^−5^) levels show large variation among batches, indicating that these variables are subject to strong technical variation.

**Figure 2.**
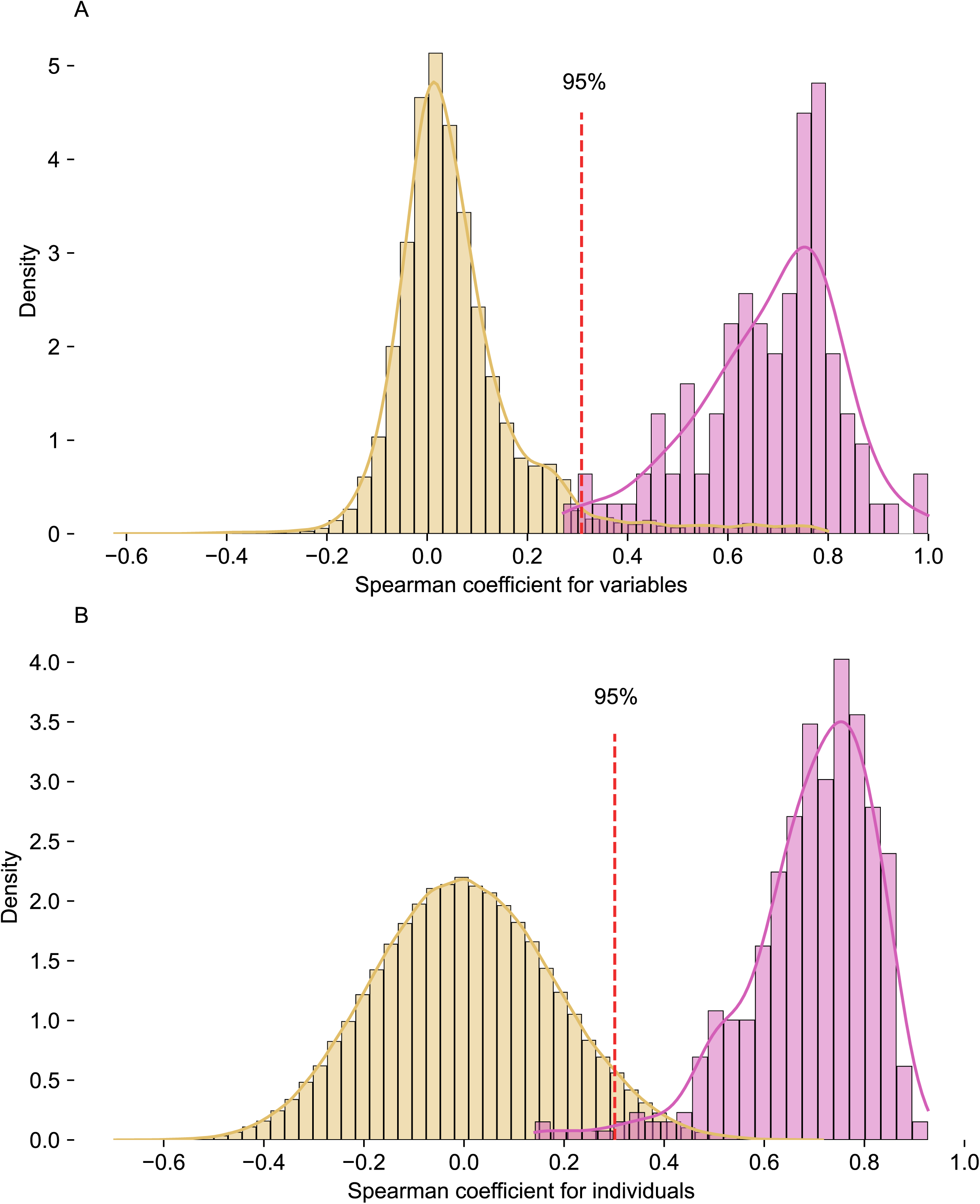
Quality of the newly collected lab measurement data in V3 visit. (Top) Density distributions of Spearman correlation coefficients between 107 unpaired (yellow) and paired (purple) variables measured in V1 and V3. The red dashed line indicates the 95^th^ percentile of the null distribution (yellow). Six individuals were discarded due to low paired correlations. (Bottom) Density distributions of Spearman correlation coefficients between 411 unpaired individuals (yellow) and paired individuals (purple) sampled in V1 and V3. The red dashed line indicates the 95^th^ percentile of the null distribution (yellow). Potassium and chlorine concentrations were discarded due to low paired correlations. (n=411)

We next assessed how age affects biological measurements by leveraging both V1 and V3 data. To discern true age effects from batch effects between V1 and V3 visits, we merged V1 and V3 data and built a model of each biological measurement with age and visit as explanatory variables. Consistent with our previous analyses, we found strong differences between visits for a few variables, including chlorine levels, bilirubin levels and basophil counts (LRT *P*_visit_ < 1.1 × 10^−14^) (Fig. S4), indicating again high measurement error. Controlling for differences between visits, we found that 39% (18/46) of variables show a significant age effect (Fig. 3). The biological measurements most associated with age, but not with visit, were fasting glycemia (β_age_= 0.013, 95% CI: [0.010 – 0.016]; *P*_age_ = 1.1 × 10^−15^), urea levels (β_age_ = 0.040, 95% CI: [0.033 – 0.047]; *P*_age_ = 5.5 × 10^−27^), and height (β_age_ = −0.11, 95% CI: [-0.059 – −0.154]; *P*_age_ = 5.4 × 10^−7^) (Fig. 3A, B, C). Several measurements showed a highly significant age effect and a significant, although low, visit effect, including systolic blood pressure (β_age_ = 0.54, 95% CI: [0.45 – 0.63]; *P*_age_ = 1.7 × 10^−30^), total cholesterol (β_age_ = 0.026, 95% CI: [0.021 – 0.032]; *P*_age_ = 1.3 × 10^−19^), and albumin (β_age_ = −0.040, 95% CI: [-0.053 – −0.028]; *P*_age_ = 1.53 x10^−10^) (Fig. 3d, E, F). However, these V1/V3 differences could also be caused by non-linear effects of age. Collectively, the high-quality longitudinal data collected on the MI cohort recapitulates well-known effects of aging on health biomarkers^36^, while accounting for the batch effects observed between visits.

**Figure 3.**
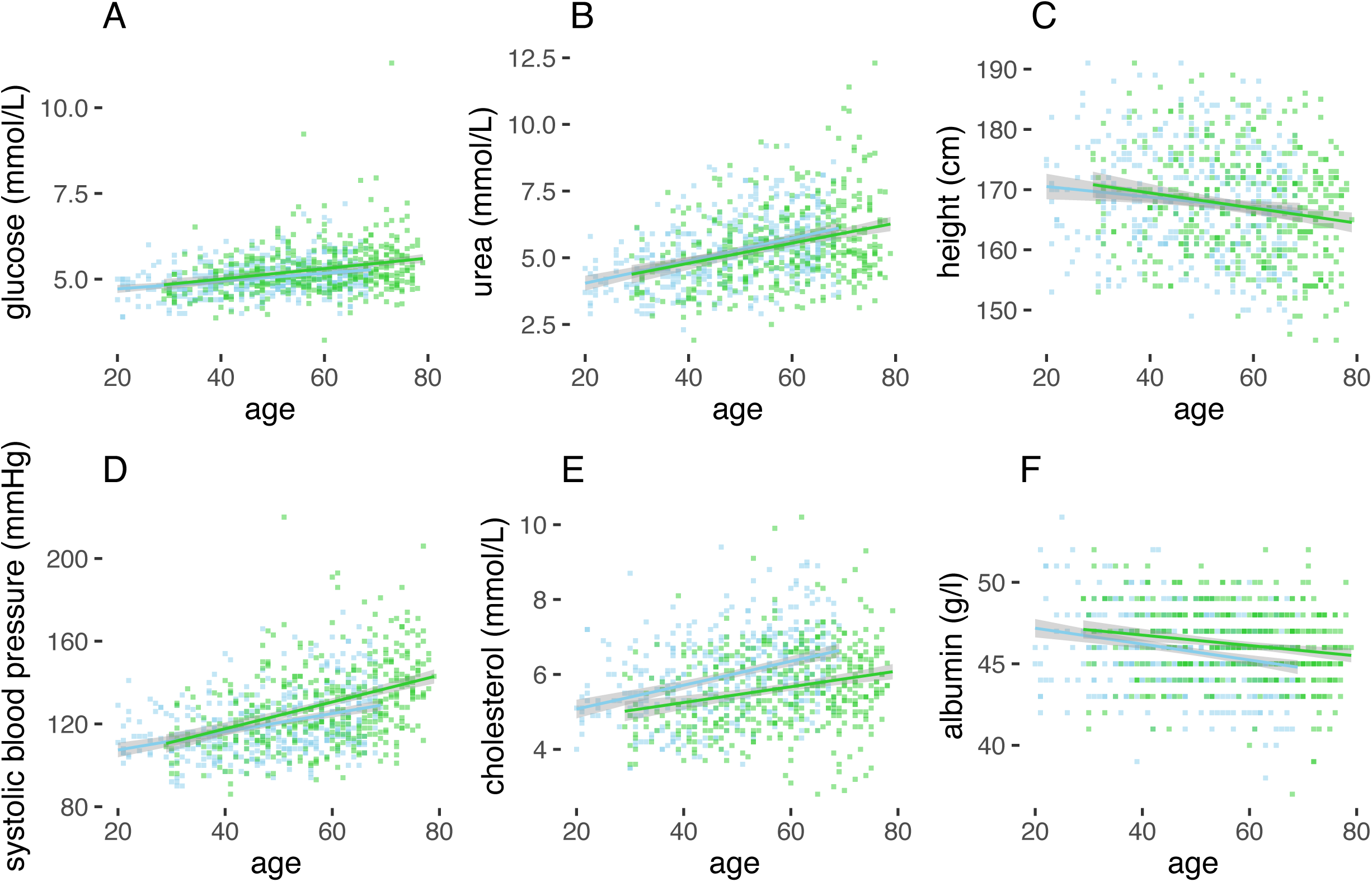
Age-related changes in health biomarkers in the Milieu Intérieur aging cohort. Scatter plots of (A) glycaemia, (B) urea concentration, (C) height, (D) systolic blood pressure, (E) total cholesterol, and (F) albumin measurements in V1 (blue) and V3 (green) visits, as a function of age. The variables most associated with age are shown. The solid straight lines indicate the linear regression line. Gray shaded areas indicate the 95% confidence intervals. (n=411)

### Large-scale serotyping and infectious history

To assess how antibody responses of MI donors changed over the 10-year period, we measured immunoglobulin levels against 72 antigens from a diverse array of viruses and bacteria responsible for vaccine-preventable diseases and common infections in plasma samples from V1 and V3. Seropositivity was determined for 45 antigens, based on previously defined cut-offs^27^, and serostatuses were compared between V1 and V3 visits (Fig. S5). We identified seroconversions for 42 out of 45 antigens. Seroconversion rates varied from 0.02% to 13.6%, with an average of only 2.6% per antigen. The antigens with the greatest number of seroconversions were, as hypothesized, those related to common infections such as seasonal coronaviruses (in particular NL63), enteroviruses, norovirus, and adenoviruses (Table S1). More surprising was the high seroconversion rate for measles (10.0%), given that this is a required childhood vaccine in France, suggesting a high incidence of natural, possibly asymptomatic, infections in the adult population^37^.

To assess age-related changes in serological measures, we next modelled the 72 available antibody levels in V1 and V3 donors as a function of age and visit. We observed an overall increase in anti-measles viral IgGs with age in donors at both time-points (Fig. 4B), suggesting continuous exposure to the measles virus over adulthood. Similar increases in antibody levels with age were observed for anti-RSV, anti-norovirus, and anti-HAV specific IgG, likely reflecting age-related differences in exposure (Fig 4A, D, E)^38,39^. In contrast, antibodies against *Bordetella pertussis* and HPV-16 remained constant with age (Fig. S6), implying sustained humoral immunity. Finally, antibodies against *Diphtheria* and the tetanus toxin both showed a decline with age at both time-points, likely reflecting waning immunity in the absence of re-exposure^40^. Interestingly, antibody levels against the diphtheria toxin (*P* = 1.91 × 10^−14^) and tetanus toxin (*P* = 6.28 × 10^−7^) were the only serological variables showing a significant non-linear effect of age (Fig. 4C, F). Both antibody titers peaked in 45-50 year-old V3 donors, perhaps reflecting different lifetime exposures to these antigens through revaccination or infection. Collectively, these results reveal dynamic changes in specific antibody responses with age in the French population and highlight the importance of re-vaccination campaigns for public health.

**Figure 4.**
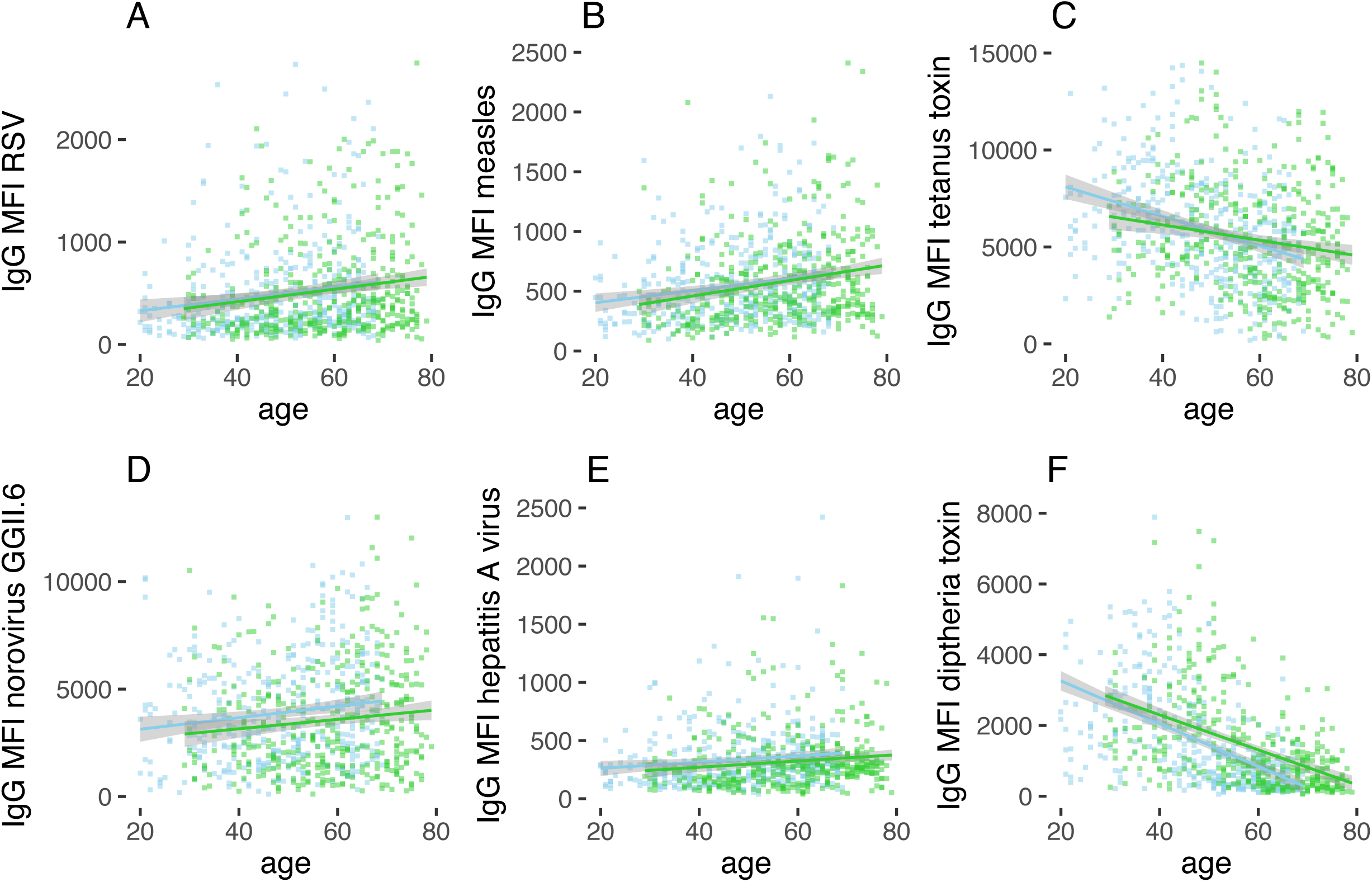
Age-related changes in antibody titers in the Milieu Intérieur aging cohort. Scatter plots of Luminex-based MFIs measuring IgGs against (A) respiratory syncytial virus (RSV), (B) measles virus, (C) tetanus toxin, (D) norovirus, (E) hepatitis A virus (HAV), and (F) diphtheria toxin in V1 (blue) and V3 (green) visits, as a function of age. (A, B, D, E) The solid straight lines indicate the linear regression line. (C, F) The solid smooth lines indicate the fitted polynomial function with three degrees of freedom. (A-F) Gray shaded areas indicate the 95% confidence intervals. (n=411)

### The impact of the COVID-19 pandemic

Given that the V3 visit occurred in 2022, close to the COVID-19 pandemic, the MI follow-up study provided an opportunity to assess immune responses to acute viral infection with SARS-CoV-2 and to COVID-19 vaccination. We included extensive questions in our eCRF related to the coronavirus infection. Among the 415 donors questioned, 32% reported a confirmed infection with SARS-CoV-2, in agreement with the reported infection rate in France during this period^41^. Confirmed SARS-CoV-2 infection was associated with younger age (LRT *P*_age_ = 4.7 × 10^−5^) and female sex (LRT *P*_sex_ = 0.02). Based on reported symptoms and treatments, half of the SARS-CoV-2 infected patients (50.4%) reported an infection severity index of 3 (Table 3), and only one patient required hospitalization (without respiratory assistance), likely reflecting the overall healthy nature of the cohort. Similarly, the proportion of donors reporting persistent symptoms consistent with long COVID was 13.3%, lower than rates reported in the general population^42^. Age, sex and six other candidate risk factors (Methods) were not significantly associated with COVID-19 severity (*P*_adj_ > 0.05), likely due to limited power.

Using serology data, we found that 15.4% (= 64/414) of participants were positive for antibodies against SARS-CoV-2 nucleoprotein (NP) (using V1 pre-pandemic samples to define the cut-off), of which 19 self-declared no infection. The resulting proportion of asymptomatic infected donors, 29.6% (= 19/64), is within the range reported in the literature^43^, though a major limitation of such studies is the poor durability of anti-NP antibodies^44^. Regarding COVID-19 vaccination, most donors reported receiving a vaccination, with only 7% (*n* = 30) not receiving any vaccine, which is lower than the estimated non-vaccination rate in France (13%). Among vaccine recipients, the vast majority (85%) received the recommended three doses, 14% received two doses, and just 1% received a single dose. Of first doses received, 70% were with the Pfizer vaccine, 15% with AztraZeneca, 9% with Moderna, and 6% with Johnson and Johnson.

### Medical events and biological aging

We examined the medical history of all V3 donors to determine whether they had experienced notable clinical events since their initial recruitment visit (V1). Out of the 415 participants, 134 reported disease occurrences, including cancer, autoimmune disease, cardiovascular disease, chronic, acute or recurrent infection, type-II diabetes, or severe allergy (**Table 2**). These donors were classified as ‘diseased’. 86 V3 donors presented risk factors for various diseases, including past/treated diseases, elevated BMI (> 32 kg/m²), hypertension, low platelet count (<120,000/mm^3^), low hemoglobin concentration (<10.0 or 11.5 g/dl for women and men, respectively), elevated transaminases, heavy alcohol consumption (> 50g of ethanol per day), or drug consumption (**Table 2**). These donors were classified as ‘at risk’, while the remaining 191 participants were classified as ‘healthy’ and would still pass the original MI V1 inclusion criteria. We found that the diseased group was significantly older than the other groups (*P* = 0.017). We next tested whether biological measurements differed among the diseased, at risk and healthy groups, excluding measurements that were used to define the groups and controlling for age and sex. We observed that no individual measure was significantly different (*P*_adj_ > 0.05) between the groups.

**Table 2.** Medical events experienced by Milieu Intérieur donors since initial recruitment.

| **Disease category** | **Number of at-risk participants** | **Details of past/treated diseases (n)** | **Number of diseased participants** | **Details of current diseases (n)** |
| --- | --- | --- | --- | --- |
| Cancers or lymphoma | 36 | Other (26)<br>Breast (8)<br>Uterus (6)<br>Lung (3)<br>Lymphatic tissue (1) | 7 | Other (3)<br>Breast (2)<br>Lung (2)<br>Lymphatic tissue (1) |
| Acquired or congenital immunodeficiencies | 7 | Immunosuppressive treatment (7) | 5 | Immunosuppressive treatment (5) |
| Autoimmune diseases | 25 | Psoriasis (19)<br>Rheumatoid arthritis (3)<br>Hashimoto (2)<br>Crohn (1) | 19 | Psoriasis (11)<br>Rheumatoid arthritis (6)<br>Hashimoto (2)<br>Vitiligo (1)<br>Sjogren (1)<br>Crohn (1) |
| Pulmonary, cardiovascular hepatic or renal diseases | 48 | Pneumonia (22)<br>Asthma (12)<br>Hypertension (6)<br>Myocardial infarction (3)<br>Chronic bronchitis (3)<br>Pulmonary embolism (2)<br>Cardiac arrhythmia 1) | 64 | Hypertension (41)<br>Asthma (15)<br>Chronic bronchitis (7)<br>Auricular fibrillation (2)<br>Cardiac arrhythmia (2)<br>Emphysema (1)<br>Pneumonia (1) |
| Neurological or convulsive problems | 4 | Epilepsy (3)<br>Peripheral neuropathy (1) | 2 | Peripheral neuropathy (2)<br>Parkinson (1) |
| Infectious diseases | - | - | 44 | Fungal infection (20)<br>Cold syndrome (10)<br>UTI (2)<br>Rhinopharyngitis (2) |
|  |  |  |  | Sinusitis (2)<br>Chickenpox (2)<br>Whooping cough (2)<br>Bronchitis (1)<br>HPV (1)<br>Lyme disease (1)<br>Tuberculosis (1)<br>Conjunctivitis (1)<br>Otitis (1) |
| Chronic bone diseases | 1 | Osteoporosis (1) | 14 | Osteoporosis (14) |
| Significant coagulation problems | 8 | Anti-coagulant treatment (8) | 1 | Anti-coagulant treatment (1) |
| Severe acute/chronic allergies | 7 | Bite allergy (3)<br>Severe allergy (2)<br>Food allergy (1)<br>Dermatitis (1) | 10 | Severe allergy (5)<br>Dermatitis (3)<br>Bite allergy (2)<br>Food allergy (1) |

**Table 3.** Clinical manifestations in SARS-CoV-2-infected Milieu Intérieur donors. Only donors with a positive PCR test were included.

| **SARS-CoV-2-infection severity index** | **Clinical manifestations** | **Sample size** |
| --- | --- | --- |
| 1 | Asymptomatic | 14 |
| 2 | Symptomatic, without treatment | 38 |
| 3 | Symptomatic, with treatment | 68 |
| 4 | Hospitalization without respiratory support | 1 |
| 5 | Hospitalization with respiratory support | 0 |

We next searched for biomarkers measured during the V1 that predict disease development in V3 10 years after. We found that none of the biological and serological variables in V1 data differed significantly between healthy, at-risk and diseased V3 donors (*P*_adj_ > 0.05). However, two aggregated scores were predictive of at-risk and disease statuses. The first score, the metabolic score^35^, measures the cumulative risk for metabolic disease and was associated with the at-risk group (*P*_at-risk_ = 6.5 × 10^−4^). This indicates that the metabolic status of V1 healthy donors predicts the development of metabolism-related risk factors with age. The second score we computed was the PhenoAge score^45^, a predictor of morbidity and mortality built upon measured levels of albumin, creatinine, glucose, alkaline phosphatase, C-reactive protein, the proportions of lymphocytes and leukocytes, mean corpuscular volume, and chronological age. We found that the PhenoAge score was strongly correlated with chronological age in both V1 and V3 cohorts (Pearson’s *R*² = 0.98 and 0.97, respectively; Fig. 5A). Biological age acceleration, defined as the difference between phenotypic and chronological age, was correlated between V1 and V3 time-points (Pearson’s *R*² = 0.38; Fig. 5B), indicating that accelerated aging is consistent over a 10-year interval. Interestingly, we observed that PhenoAge of V1 donors was significantly higher in diseased V3 donors relative to healthy donors, when controlling for chronological age (3 years difference; LRT *P* = 4.7 × 10^−3^) (Fig. 5C). Differences were more significant when comparing the V3 PhenoAge score between healthy and diseased groups (LRT *P* = 7.9 × 10^−4^) (Fig. 5D). Together, these results indicate that an aggregated score measuring accelerated biological aging is a predictor of disease development in the Milieu Intérieur cohort.

**Figure 5.**
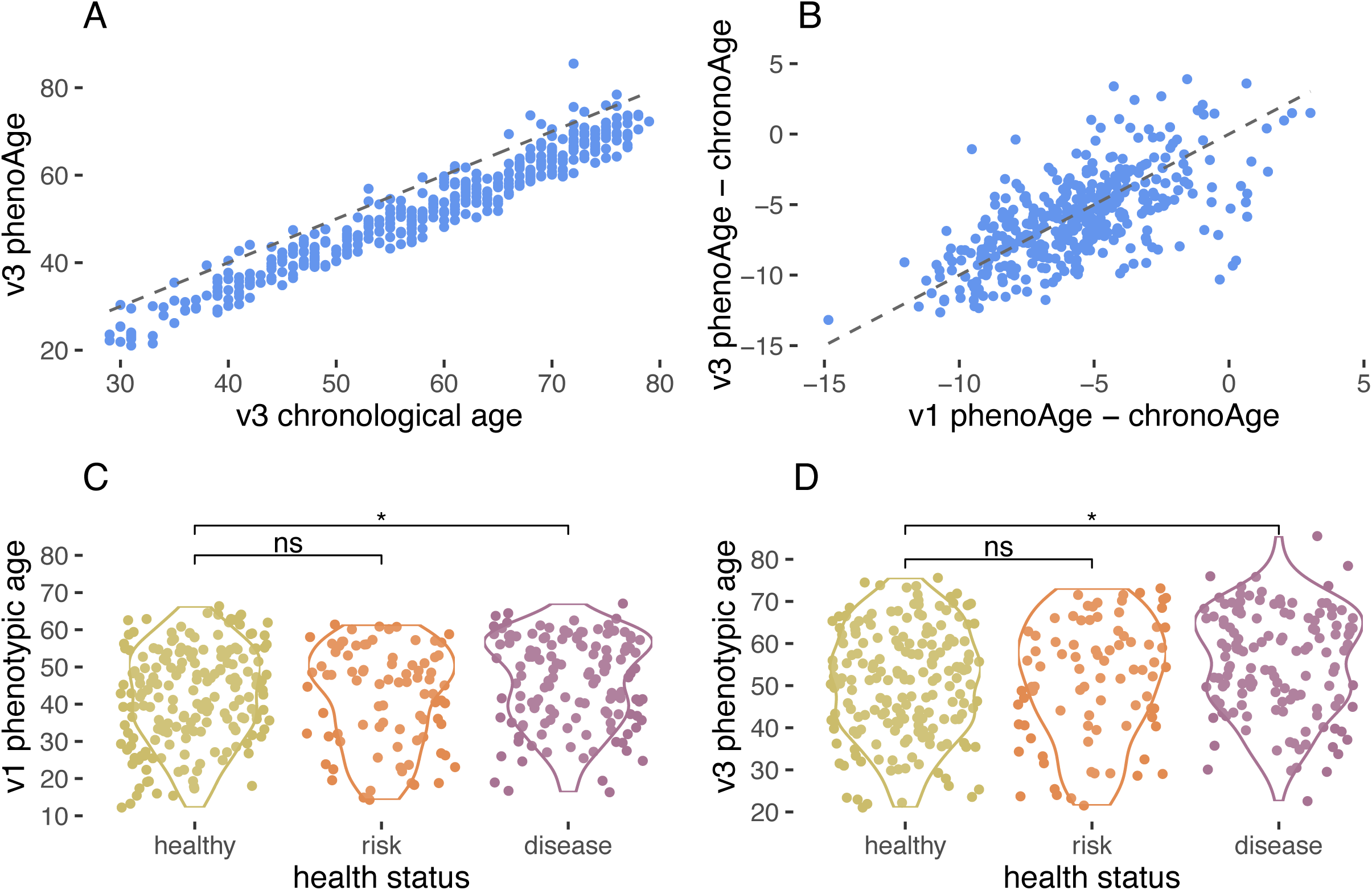
Phenotypic age and disease status in the Milieu Intérieur cohort. (A) Scatter plot of chronological age against phenotypic age in V3 visit. (B) Scatter plot of the phenotypic age acceleration in V1 visit against the phenotypic age acceleration in V3 visit. Phenotypic age acceleration refers to the difference between phenotypic and chronological age. (A, B) The gray dashed lines indicate the identity line. (C, D) Box plots of phenotypic ages estimated in (C) V1 and (D) V3 visits, as a function of disease status in V3. Asterisk and ‘ns’ indicate *P*-value < 0.05 and > 0.05, respectively, for a regression model of disease status with phenotypic age as predictor and chronological age as covariate. (n=411)

## Discussion

We present here a longitudinal study of the Milieu Intérieur cohort, which includes 415 of the original 1,000 donors sampled a decade ago. In this new V3 study, we maintained an equal sex balance but observed an overrepresentation of older donors compared to our initial sampling. Expected signs of biological aging were evident, alongside incidence of diseases within this initially healthy population. We did not observe any individual clinical laboratory measures that significantly differ between the healthy, at-risk, and diseased groups. However, a combined biological age score (PhenoAge)^45^ showed significantly elevated levels in the diseased group, highlighting the potential of biomarker-based clinical scores for accessible health monitoring^46^. Ongoing and future work will assess immune phenotypes and functions that may also differ with health status, both at this time-point and compared to data collected 10 years ago. However, the heterogeneity of diseases observed in our donors poses challenges for studies focusing on specific pathologies.

A major challenge in longitudinal studies like ours is managing technical variations in assay measurements over time. While batch effects can be corrected with statistical methods^47^, it becomes more complex when these effects are completely aligned with different groups, as can be the case across studies conducted many years apart. With this in mind, and to ensure the high-quality nature of our cohort and data set, we assessed the correlation of biological measurements between the two time-points for each donor, revealing strong correlations for the majority. For donors with lower-than-expected correlations, potential explanations include individual mismatches (which appears unlikely based on other demographic variables) or major changes over the decade. Genetic analyses will provide definitive answers, but for now, we have conservatively excluded these donors from further analyses. Reassuringly, the comparison of clinical laboratory measures also showed strong correlations for most values, with only chlorine, sodium, and albumin displaying inconsistent values.

Regarding microbial exposure assessed via serological measures, we found relatively few new infections on average over the 10-year period, although seroconversion rates for some common infections such as enterovirus and seasonal coronavirus were notably high. The obvious exception was SARS-CoV-2, for which infection rates were low and vaccination rates high in our cohort, compared to the general French population. This likely reflects an inherent bias in our cohort, consisting mostly of healthy individuals actively participating in clinical studies and well-informed about medical research. Combining self-reported infection history with extensive serological measures enabled us to identify 29% of infected individuals likely to have experienced asymptomatic infection, a group that is challenging to identify in conventional studies^43^. Further studies are ongoing to explore SARS-CoV-2-specific systemic and mucosal immunity, alongside non-specific innate immune pathways, to obtain new insight into the cellular and molecular mechanisms underlying these observations. In summary, this cohort provides a resource for studying aging immunity, with a proof-of-concept example that biological aging scores can be predictive of derivation from a healthy status. Future studies will test specific immune mechanisms that may underlie these pathological and disease conditions.

## Data Availability

All data produced in the present study are available upon reasonable request to the authors

## Acknowledgments

Milieu Intérieur is managed by the Agence Nationale de la Recherche and is supported by the French government’s Invest in the Future programme; reference ANR-10-LABX-69-01. We thank Biotrial for support with clinical study recruitment, the Institut Pasteur clinical coordination team for help with clinical study protocol management, and all the Milieu Intérieur donors for volunteering. The Milieu Intérieur Consortium¶ is composed of the following team leaders: Laurent Abel (Hôpital Necker), Andres Alcover, Hugues Aschard, Philippe Bousso, Nollaig Bourke (Trinity College Dublin), Petter Brodin (Karolinska Institutet), Pierre Bruhns, Nadine Cerf-Bensussan (INSERM UMR 1163 – Institut Imagine), Ana Cumano, Christophe D’Enfert, Ludovic Deriano, Marie-Agnès Dillies, James Di Santo, Gérard Eberl, Jost Enninga, Jacques Fellay (EPFL, Lausanne), Ivo Gomperts-Boneca, Milena Hasan, Gunilla Karlsson Hedestam (Karolinska Institutet), Serge Hercberg (Université Paris 13), Molly A Ingersoll (Institut Cochin and Institut Pasteur), Olivier Lantz (Institut Curie), Rose Anne Kenny (Trinity College Dublin), Mickaël Ménager (INSERM UMR 1163 – Institut Imagine), Frédérique Michel, Hugo Mouquet, Cliona O’Farrelly (Trinity College Dublin), Etienne Patin, Antonio Rausell (INSERM UMR 1163 – Institut Imagine), Frédéric Rieux-Laucat (INSERM UMR 1163 – Institut Imagine), Lars Rogge, Magnus Fontes (Institut Roche), Anavaj Sakuntabhai, Olivier Schwartz, Benno Schwikowski, Spencer Shorte, Frédéric Tangy, Antoine Toubert (Hôpital Saint-Louis), Mathilde Touvier (Université Paris 13), Marie-Noëlle Ungeheuer, Christophe Zimmer, Matthew L. Albert (Octant Biosciences), Darragh Duffy§, Lluis Quintana-Murci§,

¶ unless otherwise indicated, partners are located at Institut Pasteur, Paris

§ co-coordinators of the Milieu Intérieur Consortium Additional information can be found at: https://www.milieuinterieur.fr/en/

## Author Contributions

AJ performed data cleaning, data analysis, and writing and reviewing of the manuscript. FD, FR, TD, SC, CA implemented the study, collected, and processed donor samples. EV & VSA performed specific data analysis. DU & SFP provided clinical study and project management support. EB & MW generated antibody data sets. MH & VR provided technical expertise and supervised data collection. LQM, EP, & DD conceived, managed, and supervised the study and wrote the manuscript. All authors reviewed the manuscript prior to submission.

## Competing interests

The authors declare no competing interests.

## Materials & Correspondence

Requests for data sets related to this study should be addressed to darragh.duffy@pasteur and

---

**Figure S1.** Stability of quantitative measurements across Milieu Intérieur subjects between V1 and V3 visits. Spearman correlation matrix between unpaired and paired Milieu Intérieur subjects, at V1 and V3 visits. The colour gradient indicates the Spearman correlation coefficient.

**Figure S2.** Stability of quantitative measurements across laboratory and serological variables between V1 and V3 visits. Spearman correlation matrix between unpaired and paired laboratory and serological measurements, at V1 and V3 visits. The colour gradient indicates the Spearman correlation coefficient.

**Figure S3.** Batch effects on chlorine levels. (Top) Box plot of chlorine measurements as a function of the dates of sampling in the V3 visit. (Bottom) Heatmap indicating investigators per date of sampling. The colour gradient indicates the number of subjects examined per investigator, at the given date.

**Figure S4.** Age-unrelated changes in health biomarkers in the Milieu Intérieur aging cohort. Scatter plots of (A) heart rate, (B) immunoglobulin M levels, (C) log_2_-transformed C-reactive protein (CRP) levels, (D) bilirubin levels, (E) chlorine levels, and (F) log_10_-transformed basophil counts in V1 (blue) and V3 (green) visits, as a function of age. Only the variables not associated with age are shown. The solid straight lines indicate the linear regression line. Gray shaded areas indicate the 95% confidence intervals.

**Figure S5.** Seroconversion events in the Milieu Intérieur aging cohort. Scatter plots of Luminex-based MFIs measuring IgGs against 45 pathogens between V1 and V3 visits. The solid straight lines indicate the threshold between seronegativity and seropositivity. Orange and green points indicate seroreversions and seroconversions, respectively.

**Figure S6.** Age-unrelated changes in antibody titers in the Milieu Intérieur aging cohort. Scatter plots of Luminex-based MFIs measuring IgGs against (A) mumps lysate, (B) *Bordetella pertussis* toxin, (C) human papillomavirus (HPV) 16, (D) norovirus, (E) coronavirus, and (F) echovirus in V1 (blue) and V3 (green) visits, as a function of age. Only the variables not associated with age are shown. The solid straight line indicates the linear regression line. Gray shared areas indicate the 95% confidence intervals.

## Notes

### Competing Interest Statement

The authors have declared no competing interest.

### Funding Statement

Milieu Interieur is managed by the Agence Nationale de la Recherche and is supported by the French government's Invest in the Future programme; reference ANR-10-LABX-69-01.

### Author Declarations

The 10-year follow-up Milieu Interieur study, referred to as MI visit 3 (V3), was approved by the Comite de Protection des Personnes - Nord Ouest III (Committee for the protection of persons) on 27th January 2022, and by the French Agence nationale de security du medicament (ANSM) on 30th November 2011. The study was sponsored by the Institut Pasteur (ID-RCB Number: 2021-A02621-40) and conducted as a single center study without any investigational product. The protocol is registered at ClinicalTrials.gov (study# NCT05381857). As this study was designed to be a 10-year follow-up of the original Milieu Interieur V1 cohort (NCT01699893 and NCT03905993), the major inclusion criterion was previous inclusion in the V1 study.

