## Supplementary figures and images for "The *Milieu Intérieur* follow-up study – an integrative approach for studying immunological variation in an aging population"

### Supplemental Figures

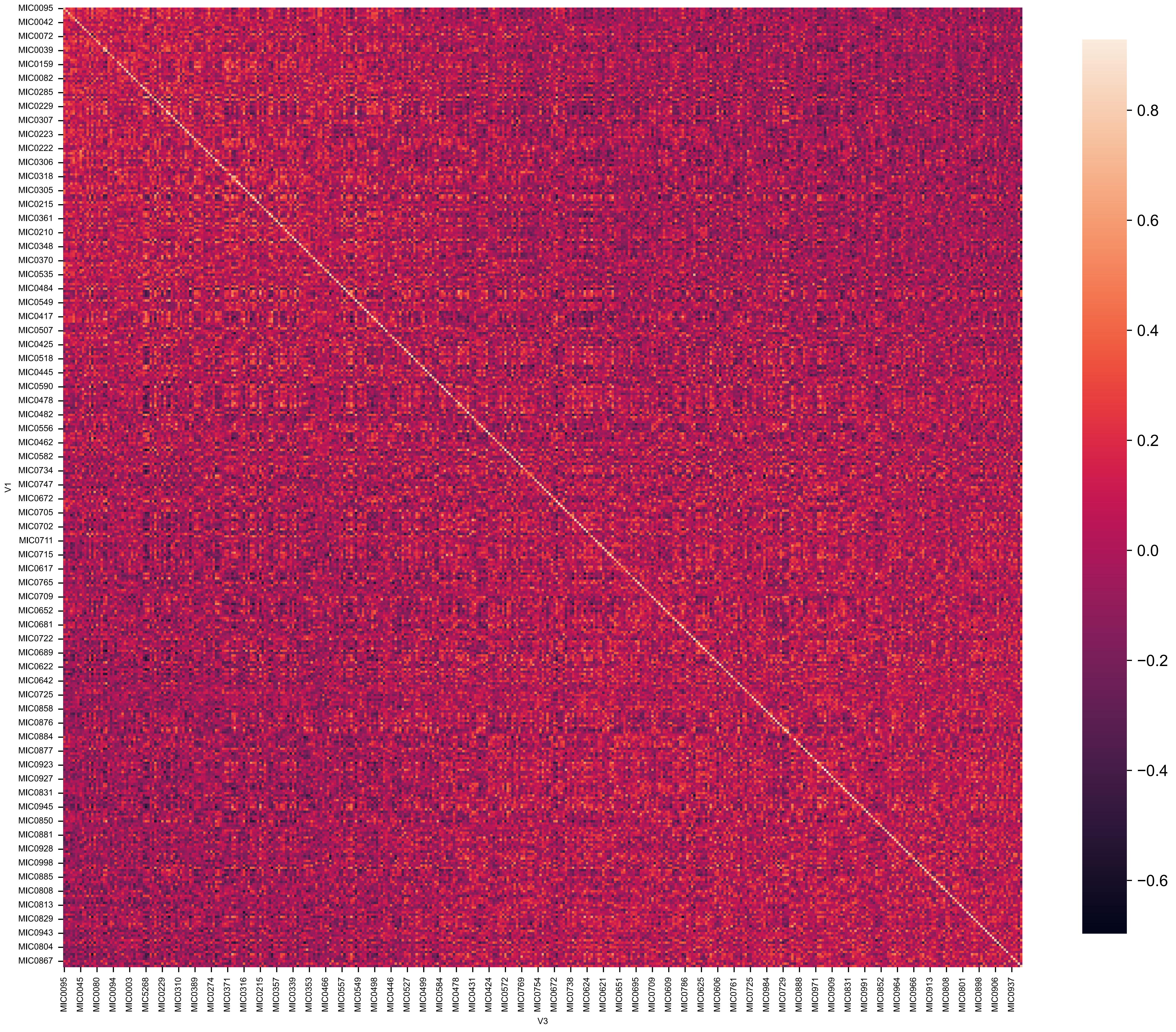

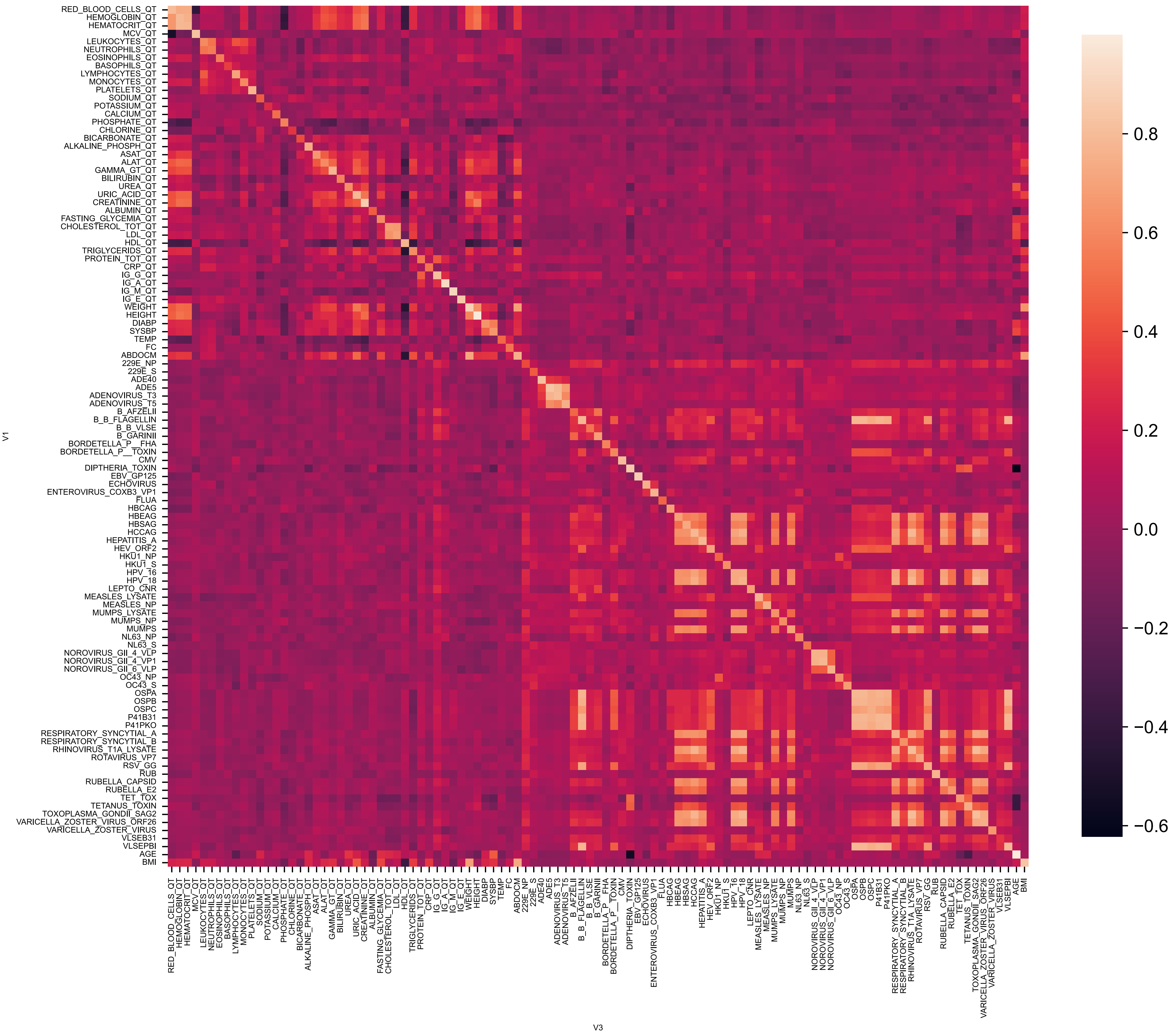

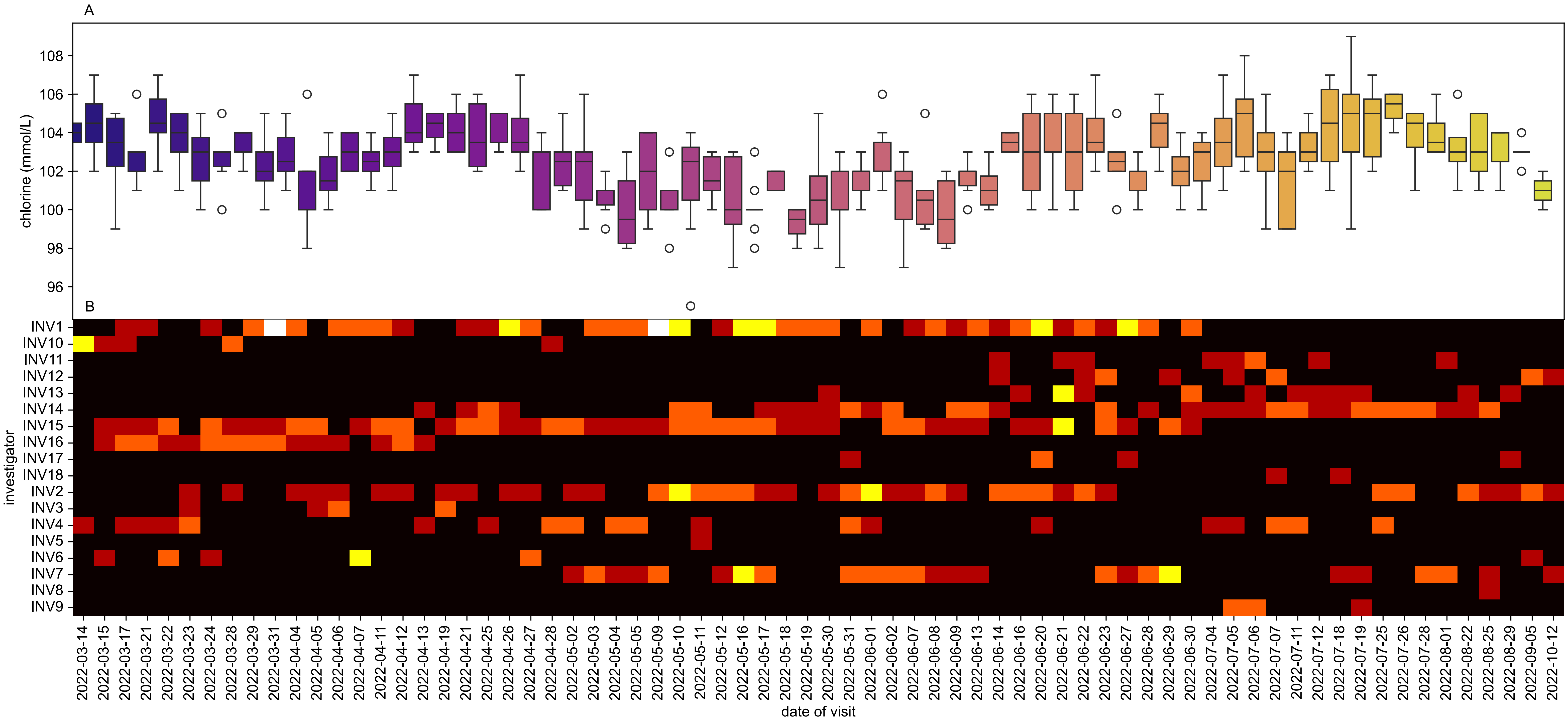

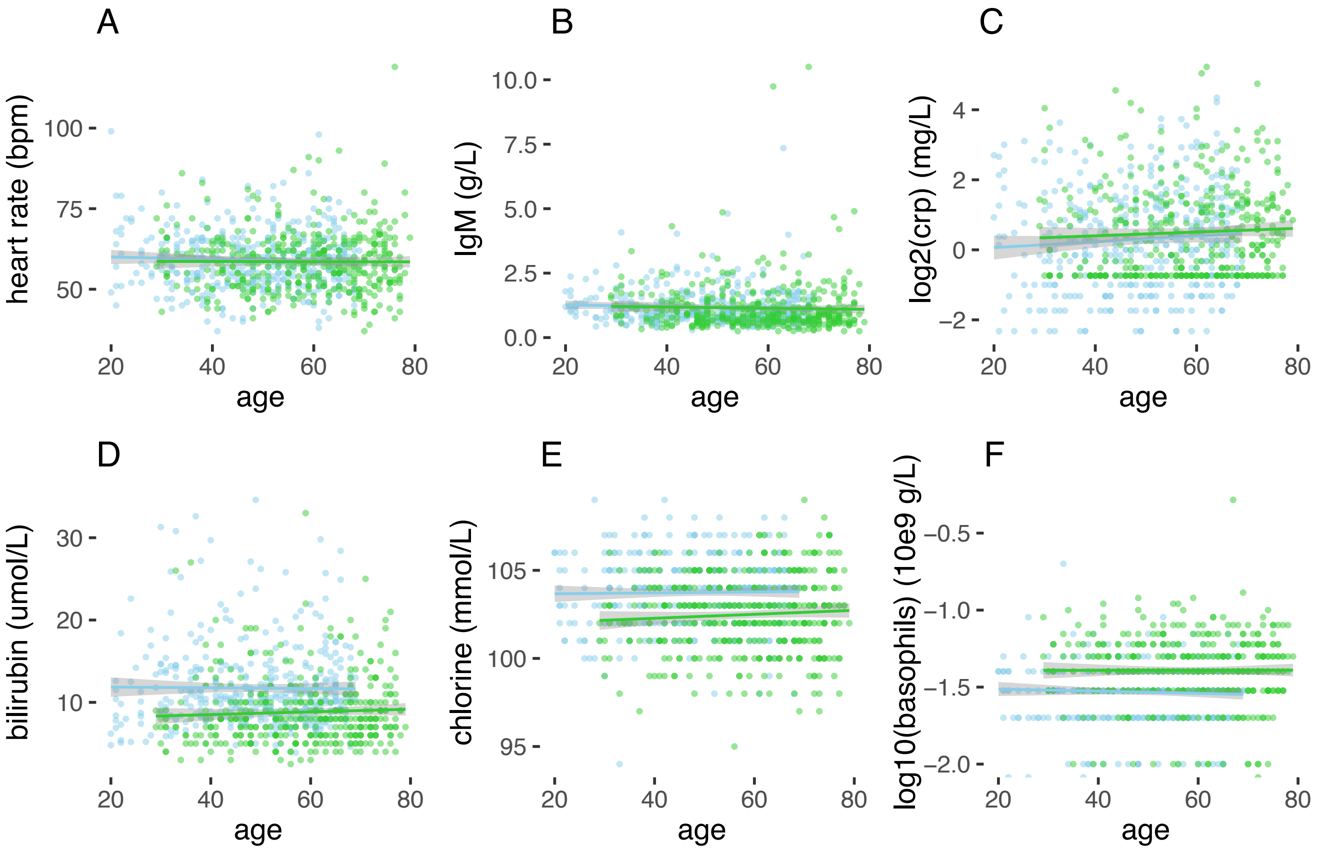

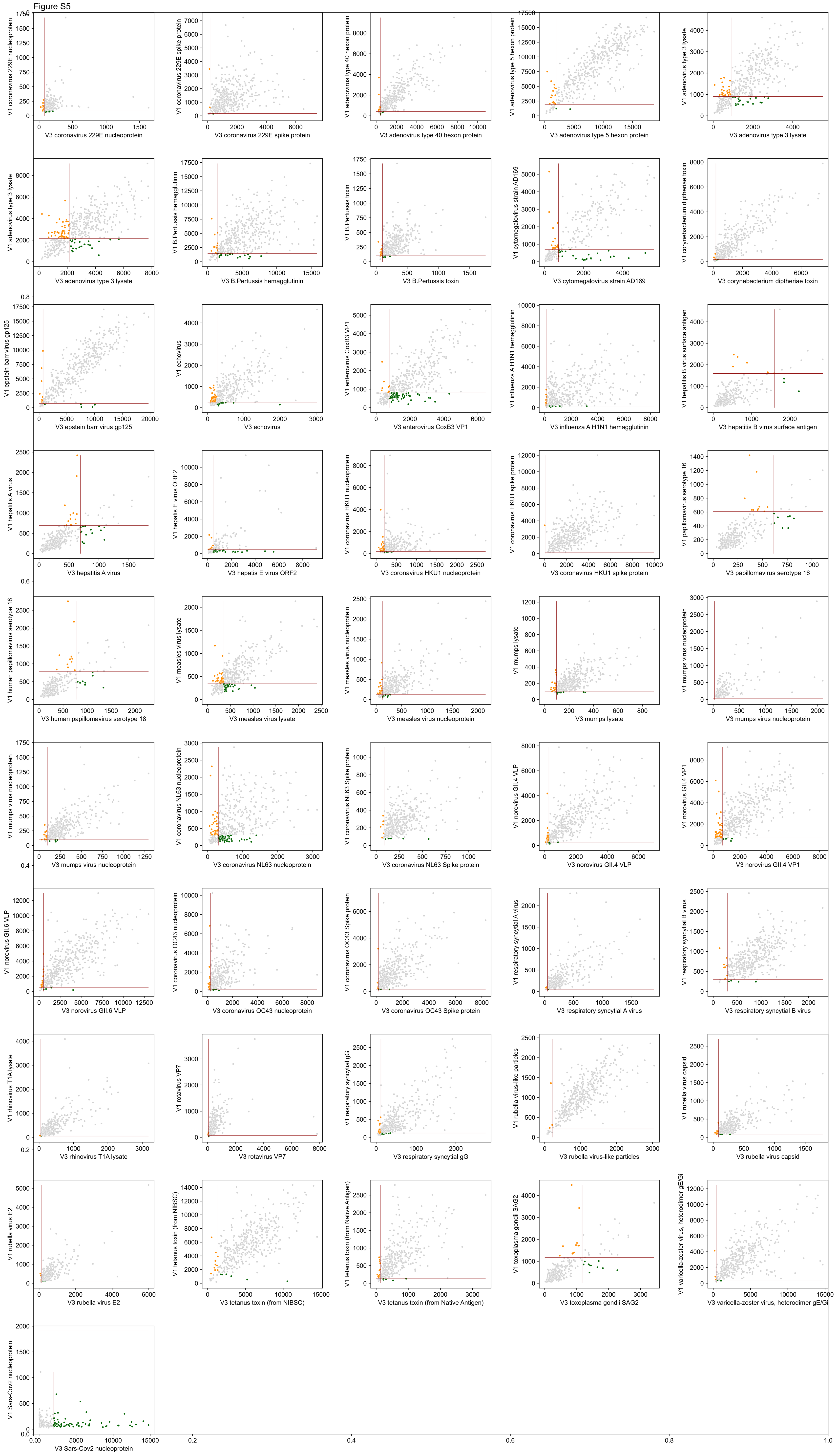

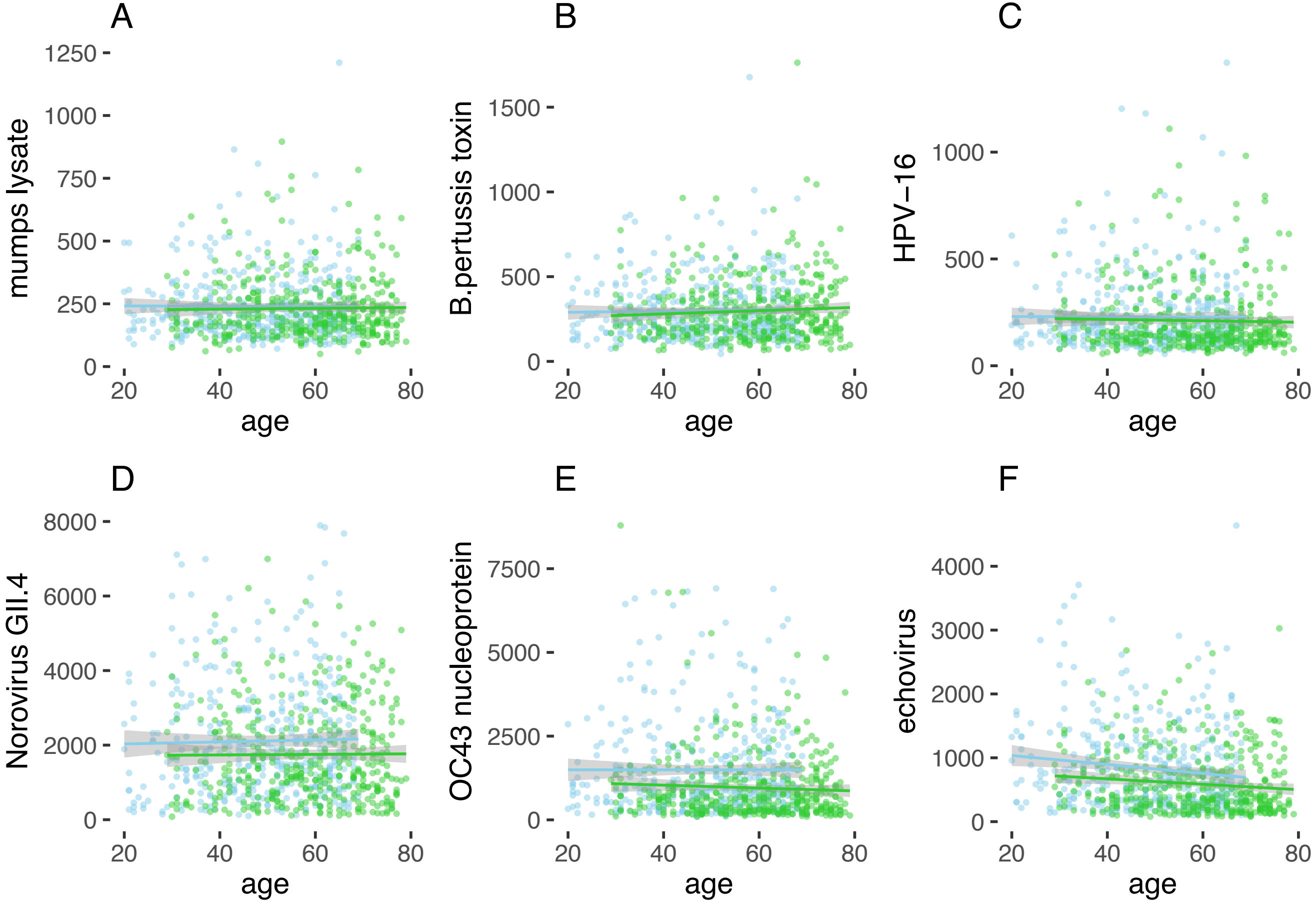
